# Population-scale integration of tumor transcriptomics into breast cancer care: a decade of the SCAN-B initiative

**DOI:** 10.64898/2026.08.20.26360879

**Authors:** Lao H. Saal, Hina Dalal, Pei Meng, Christian Brueffer, Sergii Gladchuk, Sofia K. Gruvberger-Saal, Jari Häkkinen, Nicklas Nordborg, Minerva Li, Jeanette Valcich, Ingrid Hedenfalk, Anders Edsjö, Fredrika Killander, Emma Niméus, Pär-Ola Bendahl, Carina Forsare, Jonas Manjer, Janne Malina, Martin Rehn, Kristina Åhsberg, Christian Ingvar, Fredrik Graffner, Lars Åhlund, Bengt Asking, Maria Erngrund, Monika Sjövall, Asa Cetti, Tor Svensjö, Heitti Teder, Johanna Björkman, Lena Myrskog, Anna-Karin Falck, Ann-Christine Källström, Zakaria Einbeigi, Patrícia Ribeiro Bragança, Henrik Lindman, Tobias Sjöblom, Martin Malmberg, Christer Larsson, Anna Ehinger, Lisa Rydén, Niklas Loman, Cecilia Hegardt, Åke Borg, Johan Vallon-Christersson

## Abstract

**Background:** Population-scale molecular profiling integrated into routine healthcare could accelerate biomarker discovery, validation, and implementation, but the feasibility and sustainability of such an approach have rarely been demonstrated prospectively. The Sweden Cancerome Analysis Network - Breast (SCAN-B) Initiative was established to integrate prospective molecular profiling with population-based breast cancer care and create an infrastructure for translating molecular discoveries into clinical practice (ClinicalTrials.gov identifier NCT02306096).

**Methods:** We evaluated the first 10 full calendar years of SCAN-B, encompassing patients with primary invasive breast cancer enrolled between August 30, 2010 and December 31, 2020. Enrollment and biospecimen collection were compared with all eligible breast cancer diagnoses in participating hospitals to assess population coverage and representativeness. Clinicopathological characteristics, treatments, recurrence-free survival, overall survival, RNA-sequencing-based molecular subtypes and risk-of-recurrence, and somatic mutations were evaluated. We additionally report the translation of SCAN-B molecular profiling from the research setting into routine clinical diagnostics.

**Results:** Among 16,381 estimated eligible breast cancer diagnoses, 13,940 patients (85.1%) were prospectively enrolled across participating Swedish hospitals. Baseline blood samples were obtained from 98.4% of enrolled patients and tumor specimens from 71.1%; 9,323 tumors (94.0% of submitted tumor specimens) underwent RNA-sequencing. The enrolled cohort was broadly representative of the underlying breast cancer population across major clinicopathological characteristics. Integration of longitudinal clinical data with molecular profiling enabled characterization of real-world treatment patterns, long-term outcomes, molecular subtypes, risk-of-recurrence, and the somatic mutational landscape in this population-based cohort. Building on prospective real-time RNA-sequencing and subsequent development and validation of single-sample molecular subtype and risk-of-recurrence predictors, the SCAN-B workflow was transferred into routine clinical molecular diagnostics in Skåne and Blekinge in 2021. Through January 2026, more than 3,000 patients had received clinical RNA-sequencing-based molecular subtype and risk-of-recurrence reports, while prospective SCAN-B enrollment and transfer of samples and molecular data into the research infrastructure continued. Patient enrollment continues prospectively, with over 23,000 patients accrued as of January 2026.

**Conclusions:** A prospective, population-based molecular profiling program can be integrated into routine breast cancer care at scale while maintaining high population coverage and representativeness. Over more than a decade, SCAN-B progressed from prospective biosampling and molecular profiling through biomarker development and validation to implementation of RNA sequencing-based testing in routine healthcare. This model establishes a continuous framework linking population-based molecular research, biomarker discovery and validation, and clinical implementation, and provides a strategy for integrating precision oncology research with routine cancer care.

**Trial registration:** ClinicalTrials.gov identifier NCT02306096

## INTRODUCTION

Breast cancer is one of the most common cancer diseases in Sweden and globally. In Sweden, 10,893 breast cancer (BC) diagnoses and 1395 BC-related deaths were reported in 2020. Globally, these numbers are over 2.25 million diagnoses and 680,000 deaths annually [1]. Although the incidence of breast cancer has generally increased the past 50 years, the mortality from breast cancer has been slowly decreasing. Today, the 5-year survival for breast cancer is approximately 90%. However, it is underappreciated that a 5-year survival does not mean cure for breast cancer. Overall, the rate of relapse for breast cancer is quite constant, driven primarily by the most common estrogen receptor-positive (ER+) subtypes with a roughly equal number of events occurring within 5-years as occurring after 5-years. Thus, improved prognostic and predictive tools are still of acute need to better predict who may be cured with minimal systemic therapy, and those who are at high risk for late relapses and may require additional intervention.

Molecular tests are the backbone of personalized and precision medicine. Particularly in the last 20 years, the increasing clinical use of gene expression testing has significantly transformed the management of breast cancer, providing valuable insights into tumor biology, prognosis, and treatment response [2]. One of the most widely utilized gene expression assays in breast cancer is the Oncotype DX Breast Recurrence Score^®^ (Exact Sciences), which assesses the expression levels of 21 genes involved in tumor proliferation, estrogen receptor signaling, and invasion [3]. This test aids clinicians in stratifying patients into low, intermediate, or high-risk categories, guiding treatment decisions regarding the necessity of adjuvant chemotherapy [4, 5]. Similarly, the Prosigna^®^ Breast Cancer Prognostic

Gene Signature Assay (Veracyte) evaluates the expression of 50 genes associated with breast cancer recurrence risk, assisting clinicians in prognostication and treatment planning [6].

Gene expression testing plays a pivotal role in tailoring treatment strategies for breast cancer patients, particularly those with hormone receptor-positive, HER2-negative disease [7]. By providing molecular insights into tumor behavior and aggressiveness, these tests enable clinicians to identify patients who are most likely to benefit from chemotherapy while sparing others from unnecessary treatment-related toxicity [3, 8–13]. As molecular understanding of breast cancer continues to evolve, gene expression testing holds promise for further refining risk stratification and treatment selection. Ongoing research efforts aim to identify novel gene signatures and biomarkers that can enhance the accuracy and predictive value of existing assays, ultimately optimizing patient care and outcomes [14–19]. Furthermore, the integration of gene expression data with other molecular data (such as mutations, copy number, methylation, and proteomics), clinical parameters, as well as AI-supported digital tools, is poised to revolutionize precision medicine in breast oncology, paving the way for more tailored and effective therapeutic interventions [15, 19–21].

To address these challenges in breast cancer biology and outcome prediction, in 2009 the Sweden Cancerome Analysis Network - Breast (SCAN-B) initiative was conceived [22]. SCAN-B was established with the long-term ambition to improve the management of breast cancer through comprehensive molecular profiling and data-driven research. Spearheaded by Professor Åke Borg and launched in August 2010, SCAN-B was established to leverage cutting-edge technologies such as next-generation sequencing and gene expression analysis on very large cohorts with long clinical follow-up time to be able to elucidate the complex molecular landscape of breast tumors [22–32]. By collecting extensive clinical and genomic data from thousands of early breast cancer patients prospectively and in real-time, SCAN-B seeks to unravel the heterogeneity of the disease, identify novel biomarkers for prognosis and treatment response, and ultimately, personalize therapeutic strategies for improved patient outcomes [33–43]. Herein we present an analysis of the first 10 full calendar years of patient enrollment in SCAN-B, from 30 August 2010 through 31 December 2020.

## MATERIALS AND METHODS

### Ethics Statement

This study adhered to the ethical principles outlined in the Declaration of Helsinki. Approval was granted by the Regional Ethical Review Board in Lund (registration numbers 2009/658, 2009/659, 2010/383, 2012/58, 2012/379, 2013/12, 2013/459, 2014/521, 2014/681, 2015/277, 2016/541, 2016/742, 2016/944, 2018/267), the Swedish Ethical Review Authority (registration numbers 2019-01252, 2019-00700, 2021-03552, 2024-02040-02), and the Region Skåne biobank authority and the Swedish Data Inspection Board (diary number 364-2010).

### Patients and Samples

Informed consent was obtained in written form from all participating patients after they were thoroughly informed about the study by qualified healthcare professionals. Patient enrollment in SCAN-B was from the start an integrated component of routine clinical practice, as depicted in Figure 1. Initially, the eligibility criterion encompassed those with a preoperative diagnosis of primary invasive breast cancer. However, beginning in the autumn of 2012, eligibility was expanded to include individuals with a preoperative suspicion of breast cancer, as well as patients for whom neoadjuvant therapy was a possible treatment option [22]. SCAN-B launched in 2010 at the four hospital centers in the southern region of Sweden, Skåne (Skåne University Hospital Lund, Skåne University Hospital Malmö, Central Hospital Kristianstad, and Helsingborg Hospital), as well as at Blekinge County Hospital Karlskrona, Central Hospital Växjö, and Hallands Hospital Halmstad. The sites Uppsala University Hospital joined in October 2013, Ryhov County Hospital Jönköping joined in July 2015, and the Southern Älvsborg Hospital Borås joined in February 2021. Hallands Hospital Halmstad ended inclusion in July 2021 and Jönköping ended inclusion in March 2023. In some situations, patients with no invasive disease or generalized disease at diagnosis are enrolled; analyses and enrollment statistics presented herein reflect those of patients with primary invasive early-stage breast cancer.

**Figure 1.**
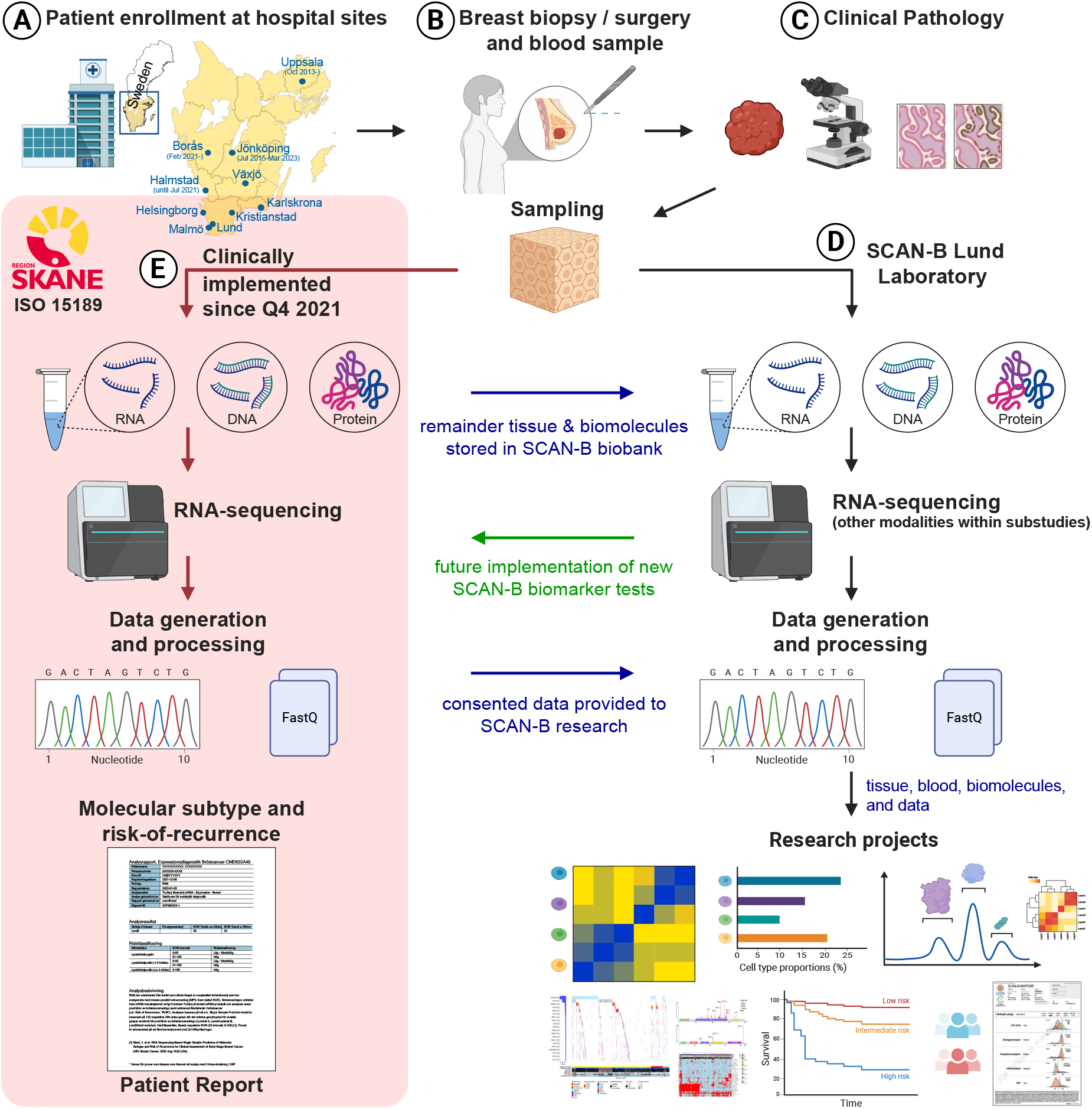
Overview of the SCAN-B infrastructure. (**A**) Patients are consented at participating SCAN-B hospital sites. (**B**) Patient undergoes surgery / biopsy per clinical routine. Baseline blood sample (and follow-up samples) are collected for research. (**C**) After routine pathological evaluation, research samples (**D**) or clinical test samples for Region Skåne (Lund, Malmö, Kristianstad, and Helsingborg) and Region Blekinge (Karlskrona) patients (**E**) are processed in the SCAN-B Lund laboratory or Section for Molecular Diagnostics Laboratory, respectively.

As SCAN-B is an observational study, all participating patients receive the same standard care afforded to non-participants. Within Region Skåne and Region Blekinge, the SCAN-B analysis for molecular subtyping and risk of recurrence [41] was transferred and translated into routine clinical practice in the autumn of 2021 and is performed for all patients in the healthcare setting within the Section for Molecular Diagnostics, Region Skåne, within the Skåne University Hospital Comprehensive Cancer Centre. This clinical translation and implementation for Region Skåne and Region Blekinge patients did not change the enrollment procedures for SCAN-B: for participating patients, the data and remaining sample derivatives are transferred to the research project (Figure 1E). Currently, for patients outside of the Region Skåne and Region Blekinge healthcare regions, SCAN-B analysis is not clinically implemented and the analyses are for research purposes only.

When patients undergo standard preoperative or pre-biopsy blood draws, an additional three 7 mL blood tubes for SCAN-B are collected and biobanked as whole blood, buffy coat, plasma, and serum (we note that plasma and serum biobanking was in place by 2012, and Jönköping and Borås sites only collect whole blood, and for all other sites, the separate aliquoting of the buffy coat ended in 2017). Clinical protocols in surgery, radiology, pathology, and oncology remain unchanged. Following pathological examination, samples of the remaining tumor specimen(s) are preserved for SCAN-B in RNAlater solution (Ambion), with preservation times carefully documented. However, after clinical implementation the Region Skåne and Region Blekinge pathology sites use a 2 mm punch biopsy to sample fresh tumor tissue from all surgical specimens where possible and preserve this in RNAlater for analysis. Not all small tumors yield study material, and clinical considerations prevent the sampling of cases found to be purely in situ carcinomas. For patients undergoing preoperative biopsy, additional study biopsies are collected and similarly preserved. Tumor sample tubes, identified by barcodes, are then transported at 4°C to the central SCAN-B research laboratory at the Division of Oncology, Lund University. Within the South Swedish healthcare region sites (Lund, Malmö, Kristianstad, Helsingborg, Halmstad, Växjö, and Karlskrona), procedures for follow-up postoperative plasma and serum sampling were implemented during 2012-2014 and are collected at 6, 12, and 36 months after the primary surgery (we note that there was an interruption during 2020-2021 due to the COVID-19 pandemic). Within subprojects of SCAN-B, additional tissue samples, blood samples, and timepoints may be collected, and after clinical implementation in Region Skåne and Region Blekinge, biobanked material remaining after the analysis is completed are transferred to the SCAN-B research laboratory on a periodic basis. All study tissues and blood materials are collected and stored centrally at the Region Skåne biobank in Lund, except for Uppsala patients whose blood samples are biobanked locally.

Clinical and pathological information linked to each patient and diagnosis, including follow-up data, is retrieved securely in electronic format on a regular basis from the National Quality Register for Breast Cancer (NKBC), housed on the Information Network for Cancer platform (INCA). NKBC is a clinical registry that systematically collects detailed data on all diagnosed breast cancer cases in Sweden, covering diagnostic procedures, tumor characteristics, treatment plans, and outcomes. Participation is nearly universal due to integration with clinical workflows and the mandate to all providers, and its completeness, timeliness, comparability and validity are exceptionally high [44]. To complement the routine flow of updates to NKBC from the clinical routine, SCAN-B has also organized focused chart reviews and updates, for example to have up-to-date follow-up information for recurrences [41].

### Processing of Tumor Specimens

In the central SCAN-B research laboratory, tumor samples preserved in RNAlater are processed as previously described [22, 30]. Each tumor is carefully weighed and, where feasible, divided into two parts: a section approximately 30 mg for concurrent DNA and RNA isolation using the Qiagen AllPrep method (see below); and any remaining tissue is cryogenically preserved for potential future analysis. For a series of the first approximately 6000 cases, an additional neighboring 10 mg section was also cut for creating a low-density, formalin-fixed paraffin-embedded (FFPE) tissue microarray (TMA). The TMA serves dual purposes, aiding in determining tumor cellularity and acts as a resource for research. Within subprojects of SCAN-B, the pathology FFPE tissue blocks for selected cohorts are collected for creation of high-density TMAs.

The AllPrep method, automated through QIAcube devices (Qiagen), is employed to extract nucleic acids and customized to preserve the protein fraction. The quality of RNA and DNA is assessed using NanoDrop spectrophotometry and either BioAnalyzer (Agilent) or Caliper LabChip XT (PerkinElmer) for capillary gel analysis. The isolation process yields RNA, DNA, and a flow-through containing proteins and short nucleic acids, all of which are biobanked for subsequent research endeavors.

To maintain the integrity and organization of study data, including sampling and analysis details, all information is meticulously documented in the SCAN-B laboratory information management system (LIMS), built upon the BioArray Software Environment (BASE) system, a secure relational data management and analysis platform originally developed for cDNA microarray sample and analysis management and later re-written and expanded with data logic layers such as ‘Reggie’ for sample management and analysis of multi-omic data [45–48]. This SCAN-B LIMS system generates intuitive sample handling and protocol workflows for laboratory personnel, ensuring consistency in laboratory operations and enhancing overall efficiency.

### RNA-Sequencing

In our central SCAN-B research laboratory, samples are processed for high-throughput RNA-sequencing (RNA-seq) largely as previously described [22, 30], with tissue processing and extractions of nucleic acids largely adhering to the original procedures whereas library preparations and sequencing has evolved, adapting to availability of kits and improved technology over time. In brief, library preparation has maintained the same overall protocol principle with poly(A) mRNA enrichment from total RNA following fragmentation and conversion into double stranded cDNA libraries with maintained RNA molecule directionality and adapter ligation with barcoded TruSeq adapters. Size-selection, PCR enrichment, and a number of clean-up steps are performed followed by quantification and quality control checks before pooling and sequencing. The original library protocol was an in-house adaption of the dUTP protocol [49, 50] implemented on a KingFisher liquid handling system. The protocol was subsequently replaced by the Illumina TruSeq stranded mRNA protocol as it became readily available. This Illumina protocol was first used as implemented on the KingFisher system, then on the Illumina NeoPrep system, and later on a Biomek4000 automated liquid handling system. Sequencing was initially done on an Illumina HiSeq 2000 with 50 bp paired-end sequencing and later replaced by Illumina NextSeq 500 with 75 bp paired-end sequencing. Single-sample prediction for breast cancer molecular subtyping (SSP-PAM50) and risk-of-recurrence (SSP-ROR) was performed as previously described [41]. Mutation calling was performed essentially as previously reported [33]. In the Region Skåne implementation of RNA-sequencing within the laboratory of the Section for Molecular Diagnostics, a similar RNA-sequencing workflow is established using the Illumina TruSeq stranded mRNA protocol implemented on a KingFisher system and sequencing on Illumina NovaSeq systems with 150 bp paired-end sequencing.

### Statistical Analysis

Analysis of enrollment and sample collection data is rooted in records maintained within the SCAN-B LIMS, with updated clinical data obtained from NKBC/INCA. Using the package ‘survival’ and ‘survminer’, survival curves were estimated using the Kaplan-Meier method and compared using the log-rank test. Recurrence-free survival (RFS) was defined as time to locoregional or distant recurrence or death, and overall survival (OS) was defined as time to death. All statistical tests were two-tailed and p-values < 0.05 were considered significant. The bioinformatics analyses conducted in this study employed an array of customized scripts in Bash, Python, and R, alongside various specialized software tools referenced above.

### Access to Data

The gene expression data derived from RNA-sequencing and microarray technologies, utilized in our prior publications, have been made publicly accessible through the NCBI Gene Expression Omnibus (GSE60789, GSE81538, GSE81540, GSE96058) and Mendeley Data repository (doi.dx.org/10.17632/yzxtxn4nmd.3) [22, 34, 41]. A complete deposit of all curated data summarized herein for the accrued SCAN-B early breast cancer cohort (2010-2020), together with RNA-sequencing gene expression data for the RNA-sequencing processed subset, is publicly available at Mendeley Data (doi.dx.org/10.17632/w8tsvd5zrf.1).

## RESULTS

### Origins and initiation of SCAN-B

In the late 1970s, Lund University’s Department of Oncology introduced the practice of analyzing hormone receptor status and S-phase fraction in fresh breast tumor tissue. This initiative significantly influenced the clinical procedures within the South Swedish Health Care Region and fostered the development of pivotal clinical trials by the South Swedish

Breast Cancer Group (SSBCG), which began in 1977. Comprising a network of professionals from various medical fields at 15 hospitals, the SSBCG aimed to equalize care standards and spearhead adjuvant therapy trials [30, 51–54]. As immunohistochemical and DNA content methods gained prominence in the 1990s [55, 56], the analysis of hormone receptors, S-phase fraction, and ploidy transitioned to pathology departments. The SSBCG remained dedicated to advancing clinical care and diagnostics, maintaining a biobank of fresh tumor samples for future research. In parallel, the Department of Oncology in the 1990s embraced molecular techniques and established a national program for hereditary BRCA gene mutation screening using Sanger sequencing methods, which transitioned to next-generation sequencing in the mid-2000s [57–61]. Around the turn-of-the-century, the Department of Oncology also was an early adopter of cDNA microarray and array comparative genomic hybridization methods, on the leading wave of transcriptional and genomic profiling applications and machine learning in breast cancer research [36, 37, 40, 62, 63].

With this rich history and experience in biomarker test development and implementation, the engagement of cooperative groups, and success in pivotal clinical trials, the idea for a large breast cancer network to develop the next-generation of diagnostic assays using the next-generation of sequencing methods was sparked in 2009. The SCAN-B initiative is a collaborative effort encompassing a broad network that includes researchers, medical professionals, and clinical staff across Sweden. This collaboration is centered at the Division of Oncology at Lund University and extends to all hospitals within the South Sweden Breast Cancer Group, with support from the Regional Cancer Center South, and the healthcare providers across 7 hospital centers within the Southern Healthcare Region, namely Malmö, Lund, Helsingborg, Kristianstad, Halmstad, Växjö, and Karlskrona. Later, additional sites at Uppsala, Jönköping, and Borås joined the SCAN-B network in 2013, 2015, and 2021, respectively. The organizational structure and partnerships that constitute the study’s foundation are detailed in Figure 1.

### Population-based Enrollment

We summarize patient enrollment for the first 10 full calendar years of the SCAN-B study, for the period from first patient enrolled on 30 August 2010 through 31 December 2020.

Patients with breast cancer are being enrolled on a rolling basis at a rate of 25 to 30 individuals per week. During this timeframe, a total of 13,940 patients (13,853 women and 87 men) with primary invasive breast cancer were consented and enrolled in SCAN-B (Table 1; Figure 2). To estimate the total number of breast cancer diagnoses meeting SCAN-B eligibility criteria, we utilized the national NKBC/INCA registry filtered by hospital site and dates of each sites’ participation: the “background” of patients diagnosed with invasive primary breast cancer patients is estimated to be 16,381 patients (Figure 2). Due to a number of factors including referral of patients between hospitals, migration of patients in/out of the catchment area of participating sites, and other factors, the total background population of eligible patients can only be approximated. From within this background, the 13,940 patients consented and enrolled in SCAN-B represents approximately 85.1% of eligible patients in the catchment area (Figure 2). From this accrued group, 13,711 have provided a baseline blood sample (98.4%), and from 9,915 patients (71.1%) a tumor tissue sample was available after pathology routines for SCAN-B research. For 9,323 of these cases, curated RNA-seq data have been generated, reflecting the extensive breadth and depth of the research cohort (Figure 2). Compared to the eligible background, this corresponds to 60.5% accrued with a tumor specimen, 24.7% accrued but without a tumor specimen, and 14.9% not accrued. The primary reason for non-accrual is language or other barrier to the informed consent process, or simply not being asked. The accrued SCAN-B cohort is generally similar in clinicopathological characteristics to the background across most variables such as estrogen receptor (ER) status, progesterone receptor (PgR) status, human epidermal growth factor receptor 2 (HER2) status, age of the patient, Nottingham histologic grade (NHG), tumor size, lymph node status, histological type, type of surgery, clinical biomarker group, and therapies received (Table 1; Figure 3).

**Figure 2.**
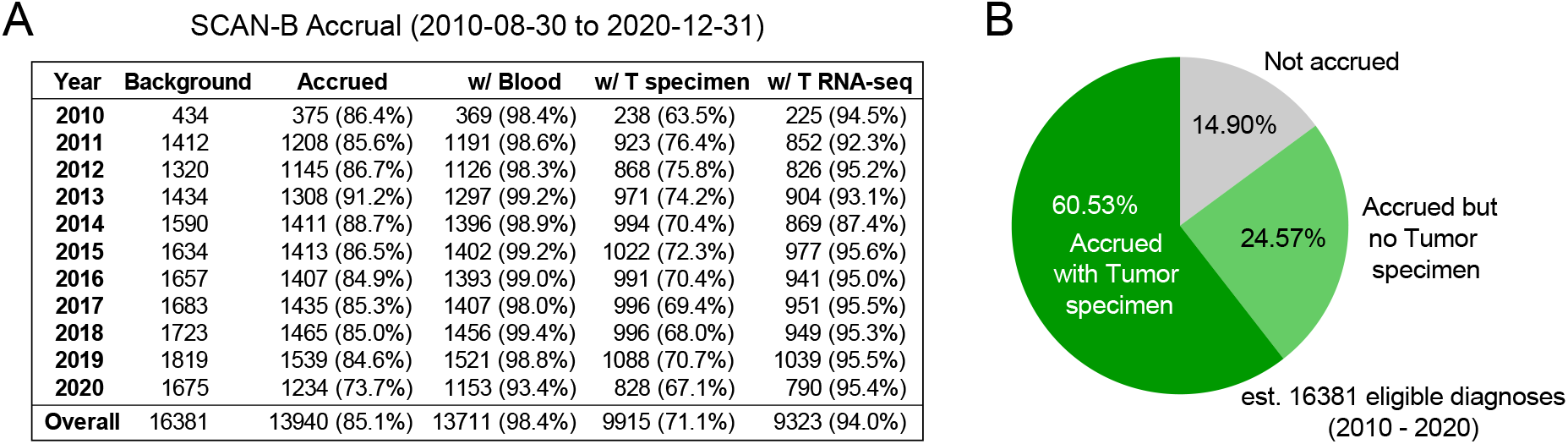
Patient enrollment. (**A**) Yearly enrollment statistics since initiation of SCAN-B from end of August 2010 and including 10 full calendar years 2011 through the end of 2020. For each year, the ‘Background’ indicates the total number of eligible diagnoses in the catchment area, followed by columns for the patients consented (‘Accrued’), those accrued and with at least one blood sample collected (‘w/ Blood’), those accrued and with at least one tumor specimen (‘w/ T specimen’), and accrued with at least one tumor RNA-seq analyzed (‘w/ T RNA-seq’). T = tumor. (**B**) Summary pie chart illustrating the proportion of all eligible diagnoses that were consented and accrued into SCAN-B but with no tumor specimen submitted, those consented and accrued and with a tumor specimen, and those who were not consented and accrued. est. = estimated.

**Figure 3.**
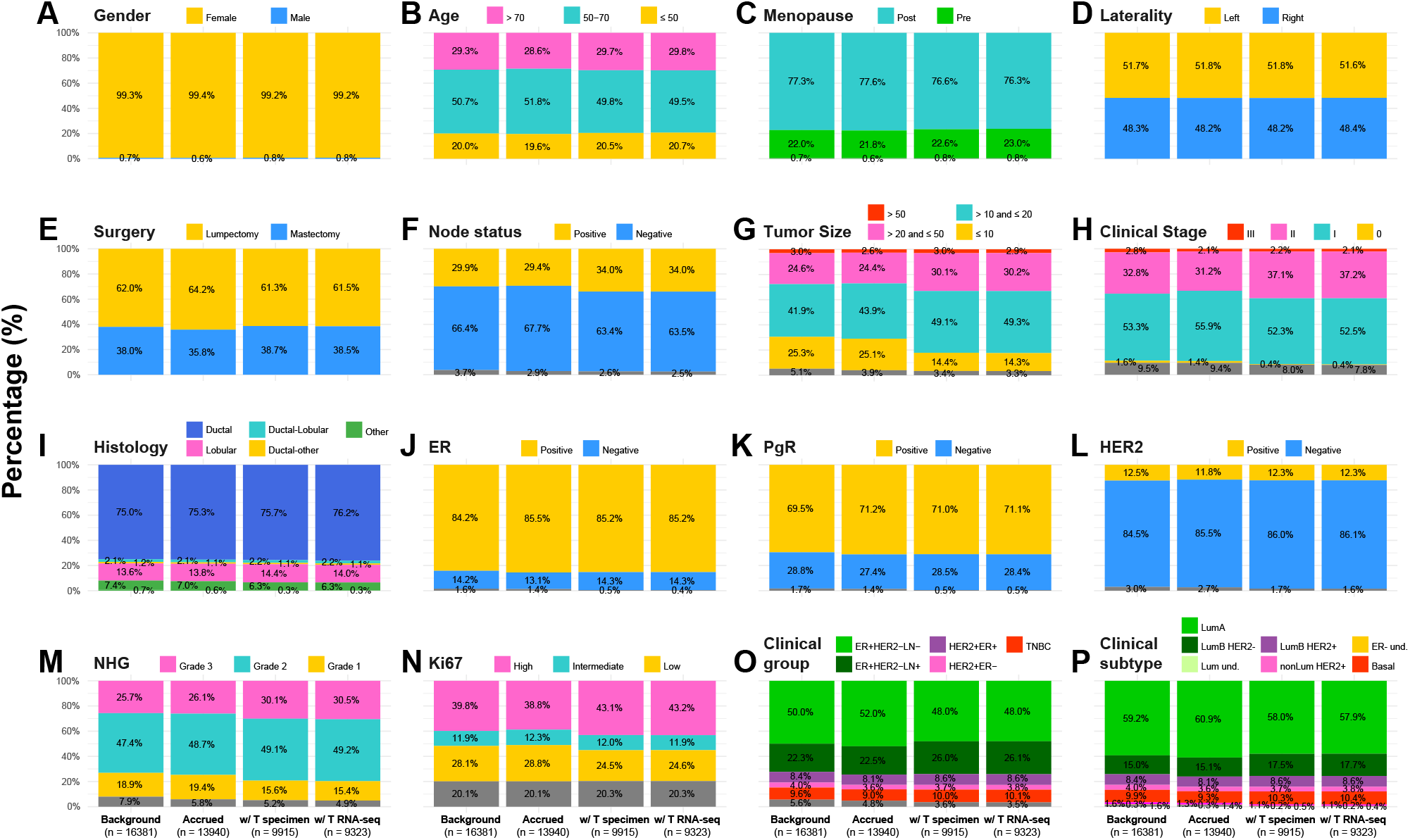
Biomarker distributions. For each biomarker, percentage distributions are provided for the four stacked groups indicated at the bottom of the figure: across all diagnoses in the catchment area (‘Background’), those consented (‘Accrued’), those accrued and with at least one tumor specimen (‘w/ T specimen’), and accrued with at least one tumor RNA-seq analyzed (‘w/ T RNA-seq’). The total n in each group is provided in parentheses. ER = estrogen receptor; PgR = progesterone receptor; NHG = Nottingham histological grade.

**Table 1:**
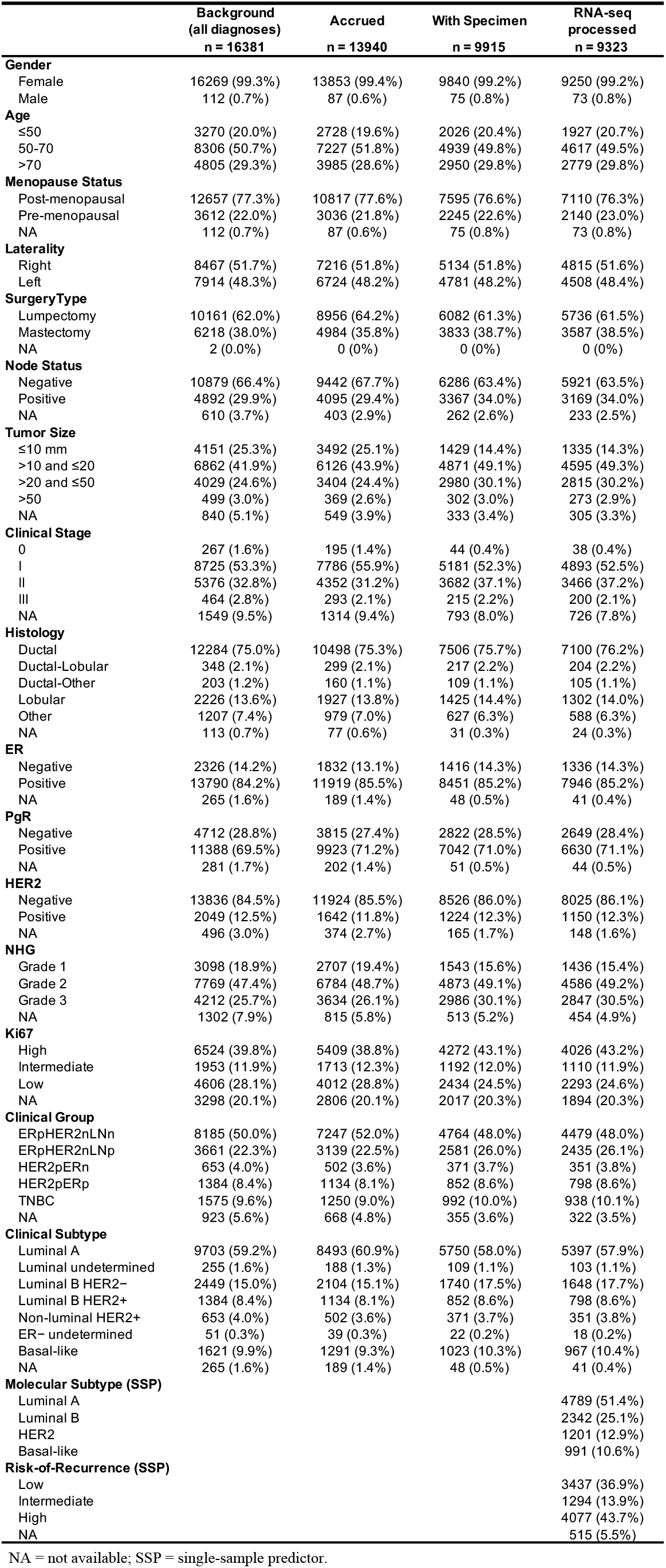
Patient demographics and clinicopathological information.

For the 71.1% of enrolled patients for which at least one tumor sample has been forwarded to our central lab, these tissue samples typically arrive 1 to 3 days after the date of biopsy/surgery. The primary reasons for not forwarding a tumor sample include when the pathologist deems the material too small for sampling or identifies the tumor as solely carcinoma in situ. The subset of patients from whom tumor samples were obtained is generally similar to the enrolled patient group, with the exception of node status, tumor size, clinical stage, NHG, and Ki67, which likely are a reflection of the undersampling of the smallest tumors (Figure 3). The median time for tumor specimens to be preserved post-removal is 55 minutes, with an interquartile range (IQR) of 38 to 80 minutes, and the median weight of these specimens is 53.1 mg (IQR, 28.8 to 95.6 mg).

In our central SCAN-B laboratory, tumor samples are processed upon receipt, divided for AllPrep and reserve pieces, processed, with RNA, DNA, and flow-through (containing proteins and small RNAs) fractions separated (see Methods), and the RNA-sequencing performed on a rolling basis. The median quantity of isolated RNA is 6.2 µg (IQR, 2.6 to 12.4 µg) and DNA is 11.4 µg (IQR, 5.3 to 20.5 µg), averaging a yield of approximately 0.6 µg RNA per mg tumor tissue and 1.0 µg DNA per mg tumor tissue. The RNA quality is generally high, with median RNA quality score (RQS) or RNA integrity score (RIN) of 8.4 (IQR, 7.8 to 8.8); DNA quality metrics are not routinely measured.

### Clinicopathological Variables

This 10-year focused overview of SCAN-B provides the opportunity to review the natural distribution of clinicopathological variables representative of Swedish patients with invasive primary breast cancer. The majority of patients are diagnosed over the age of 50, post-menopausal, with nearly 30% over 70 years and approximately 20% aged 50 or younger (see columns for ‘Background’ in Table 1 and Figure 3). Lumpectomy (partial mastectomy) is performed most frequently, approximately 30% of patients are lymph node positive at diagnosis, and about half of tumors are ≤ 20 mm (T1) and clinical Stage I. About three-fourths of cases are ductal histological type, ∼84% are ER-positive, ∼70% are PgR-positive, ∼12.5% are HER2-positive, almost a half are Grade 2 and a quarter are Grade 3, and nearly two of five are Ki67-high. In terms of clinical groups, half of cases are ER-positive, HER2-negative, and lymph node-negative (ER+ HER2− LN−) and almost 10% are triple-negative (TNBC). Utilizing clinicopathological variables for surrogate breast cancer subtyping, nearly 60% of cases are luminal A, 15% are luminal B HER2−, 8.4% are luminal B HER2+, 4% are non-luminal HER2+, 9.9% are triple-negative basal-like (TNBC), and 3.5% have no specific subtype definable due to missing information. As seen in Table 1 and Figure 3, the distribution of clinicopathological variables for patients enrolled in SCAN-B is quite consistent with the background of all diagnoses in the catchment areas, indicating the general representative nature of the SCAN-B cohort.

### Analysis of Treatment Profiles

Sweden has a universal healthcare system and nationalized treatment guidelines for breast cancer are established and updated yearly. With a focus on the SCAN-B cohort, an overview of the treatments administered to the 13,940 enrolled patients is shown in Figure 4, categorized into pre- (neoadjuvant) and post-surgical (adjuvant) therapeutic interventions.

**Figure 4.**
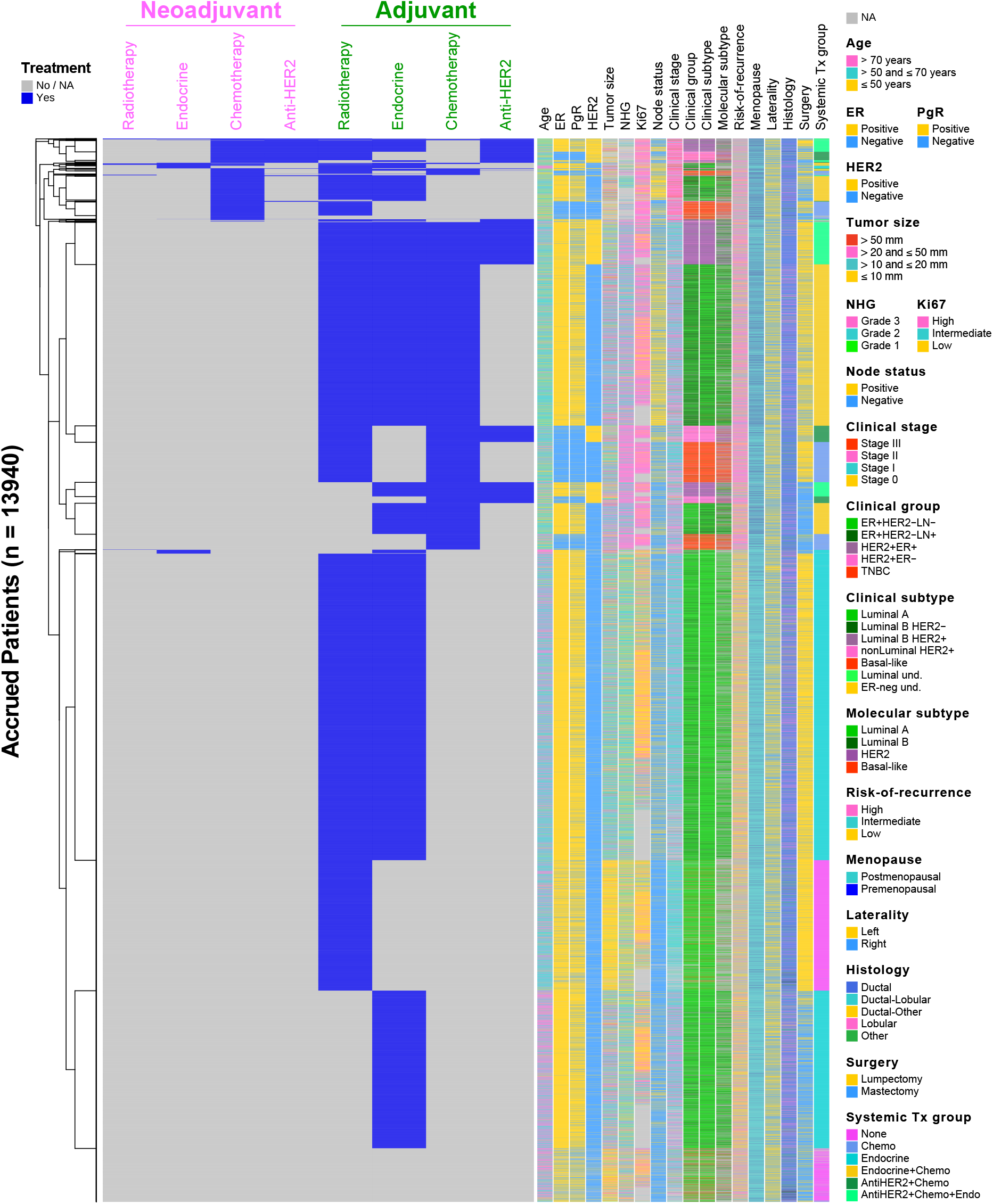
Real world breast cancer therapeutic pathways. Across the 13,940 patients enrolled in SCAN-B during the reported period (rows), the pre-operative treatments and post-operative treatments (indicated in columns) are provided. Patients are clustered according to treatments. Clinicopathological annotations and key are provided to the right.

The patients are clustered according to the treatment data. A number of observations are valuable to point out. Approximately 5.0% of patients received no neoadjuvant nor adjuvant systemic therapy, and 12.3% of patients only received radiation therapy. Overall, 8.1% of patients received at least one systemic neoadjuvant therapy: 7.5% received chemotherapy, 2.5% anti-HER2 therapy, and 1.0% endocrine therapy. Treatment patterns were time-dependent, with approximately 2.9% of patients receiving these neoadjuvant therapies in 2010 and 2011, increasing to approximately 13.0% of patients receiving these in the years 2019-2020. Post-operatively, overall, 72.5% of patients received radiotherapy, 72.1% received adjuvant endocrine therapy, 31.6% received adjuvant chemotherapy, and 10.1% received adjuvant anti-HER2 treatment. In general, post-operative treatment of patients has increased over this 10-year SCAN-B cohort duration: radiation therapy increased 1.3-fold from approximately 58.8% in 2010-2011 to 76.9% in 2019-2020; endocrine therapy increased 1.09-fold from 69.1% to 75.3%, respectively; chemotherapy usage increased by 1.19-fold from 26.2% to 31.3%; and anti-HER2 therapy by 1.18-fold from 9.0% to 10.7%.

As most patients are post-menopausal, an aromatase inhibitor (AI) is the most common type of endocrine therapy and is prescribed 1.56-fold more often than tamoxifen (TAM): of cases receiving endocrine therapy, 63.0% receive AI and 40.5% receive TAM. As for chemotherapy, 67.7% of patients receiving chemotherapy receive both a taxane and anthracycline, whereas 27.3% receive only an anthracycline-containing treatment.

### Survival Analyses

A comprehensive survival analysis for various clinical subgroups within the SCAN-B cohort was performed (Figure 5), encompassing a total of 13,938 patients. Across all SCAN-B breast cancer patients, the 5-year recurrence-free survival (RFS) was 89.1% and 76.8% at 10-years (Figure 5A), and the overall survival was 91.6% at 5-years and 80.1% at 10-years (Figure 5B). We next investigated long-term outcomes for five key clinical subgroups: ER+ and HER2– and LN– (ER+HER2−LN−), ER+ and HER2− and LN+ (ER+HER2−LN+), HER2+ and ER+ (HER2+ER+), HER2+and ER− (HER2+ER−), and TNBC. As expected, ER+HER2−LN− have the most favorable outcomes, with approximately 92.4% RFS at 5-years and 79.8% RFS at 10-years, and 94.3% OS at 5-years and 83.4% OS at 10-years (Figure 5C-D). Conversely, TNBC have the worst prognosis, with 78.9% 5-year RFS, 66.4% 10-year RFS, and 83.0% 5-year OS and 69.8% 10-year OS. The HER2 positive subgroups have a slightly worse outcome at 5-years than ER+HER2−LN−, but in the long term tend to show favorable outcomes on par with ER+HER2−LN− by 10-years after diagnosis, reflecting the impact of targeted HER2 therapies. A comparable analysis, by molecular subtype, for SCAN-B patients with tumor specimen and RNA-seq data, is provided in Supplementary Figure 1.

**Figure 5.**
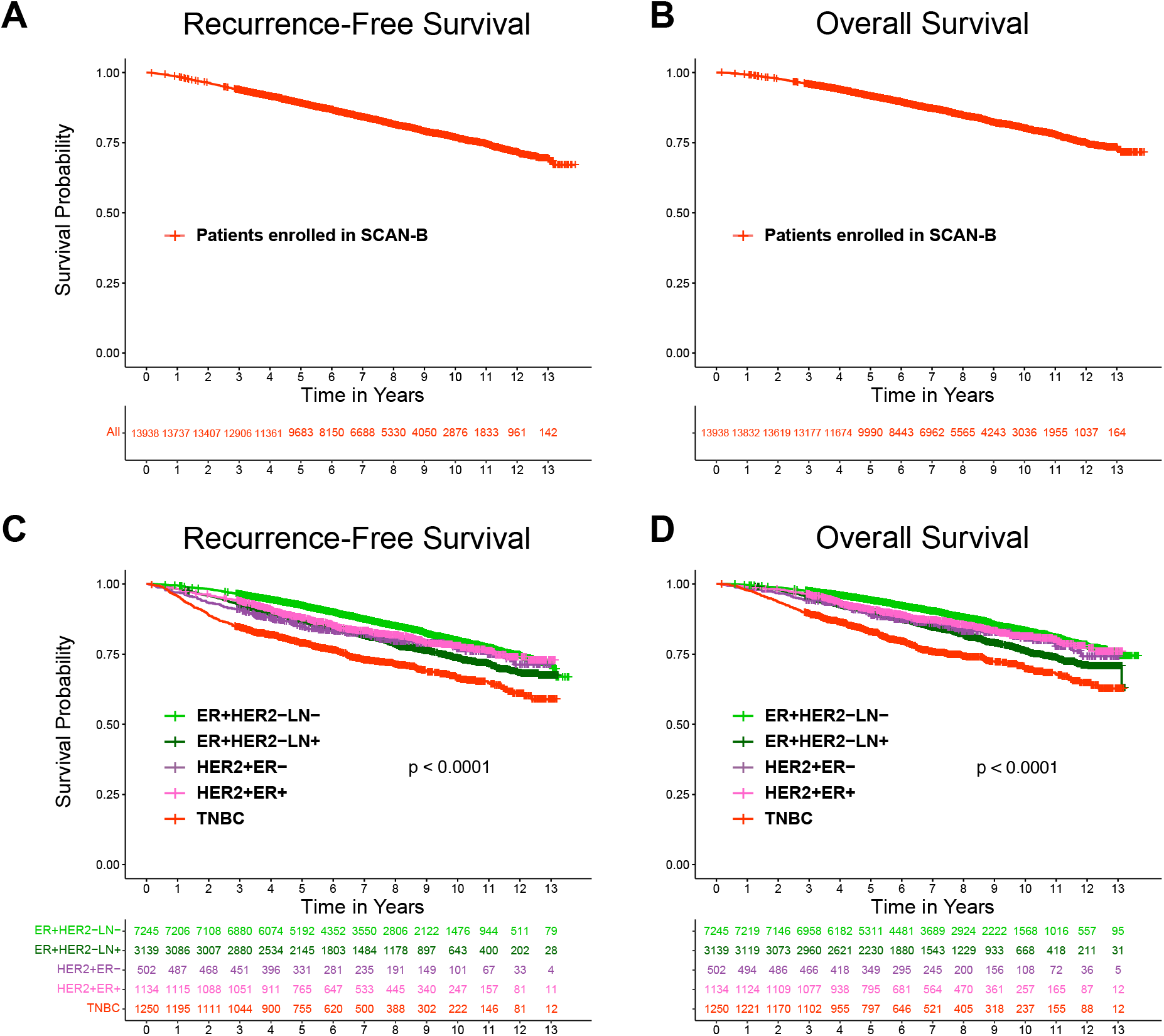
Analyses of all SCAN-B patient outcomes and by clinical groups. Kaplan-Meier survival estimates for all accrued SCAN-B patients within the reported period and with available follow-up information for recurrence-free survival (**A**) and overall survival (**B**). When patients are subdivided by clinical group, the groups have significantly different (**C**) recurrence-free survival and (**D**) overall survival.

Figure 6 provides the Kaplan-Meier survival plots of SCAN-B ER+HER2− patients who received various endocrine therapies, chemotherapies, or both. As illustrated, ER+HER2−LN+ tumors in which estrogenic signaling was not targeted (either no therapy or chemotherapy only) had particularly poor RFS and OS, whereas for LN− cases this was not as apparent: patients receiving no therapy had slightly better OS than patients receiving endocrine-only, which is likely because most of these patients receiving no therapy were clinical stage 1, whereas those receiving endocrine-only were generally of higher stages. This comprehensive survival analysis effectively showcases the critical importance of endocrine treatments in improving survival outcomes for ER+ breast cancer patients. In a similar way, anti-HER2 treatment has revolutionized outcomes for patients with amplification and overexpression of HER2. Patients who do not receive anti-HER2 targeting antibodies have a dismal RFS and OS (66.1% RFS at 5-years and 50.9% at 10-years, 70.2% OS at 5-years and 54.3% at 10-years; Figure 7A-B). The same is true for chemotherapy for TNBC: patients who do not receive any chemotherapy for TNBC has a rapid progression (63.2% RFS at 5-years and 41.9% at 10-years) and death (67.2% OS at 5-years and 44.9% at 10-years; Figure 7C-D). Observe that the data on survival outcomes in relation to treatment is non-randomized and that the different groups are not comparable in terms of risk factors, comorbidities, and age. For example, omission of systemic therapies for HER2+ and TNBC is most often due to comorbidities and age, with 56.4% and 69.6% of those not receiving systemic therapy being over the age of 70, respectively, compared to approximately 16% being over 70 years in the HER2+ and TNBC groups that did receive these therapies.

**Figure 6.**
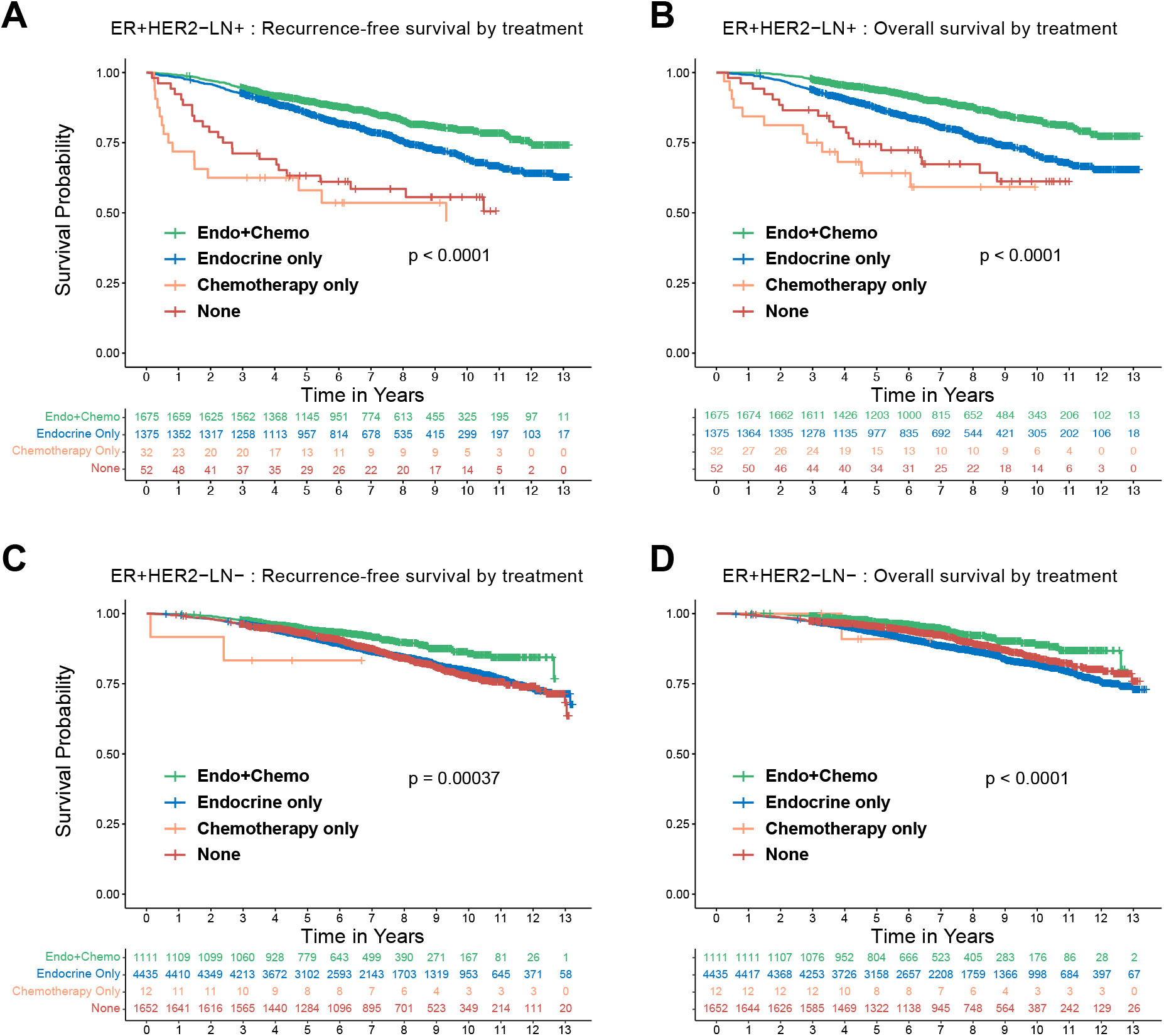
Analyses of patient outcomes within ER-positive HER2-negative breast cancer. ER-positive HER2-negative LN-positive cases have significantly different (**A**) recurrence-free survival and (**B**) overall survival dependent on whether endocrine therapy, chemotherapy, neither, or both (Endo+Chemo) is received. ER-positive HER2-negative LN-negative cases have significantly different (**C**) recurrence-free survival and (**D**) overall survival dependent on therapy received.

**Figure 7.**
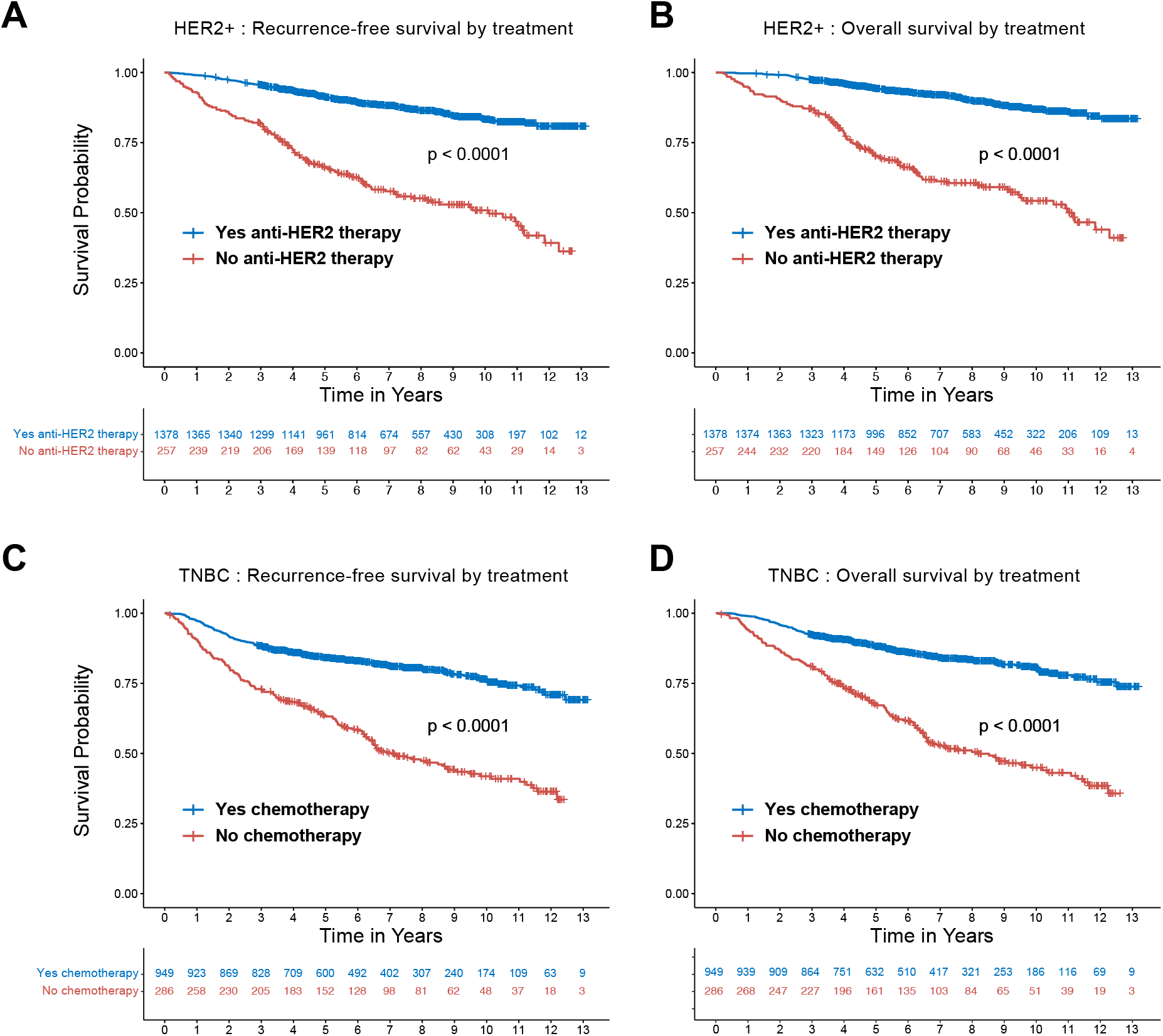
Analyses of patient outcomes within HER2-positive and triple-negative breast cancer. HER2-positive cases have significantly different (**A**) recurrence-free survival and (**B**) overall survival dependent on whether anti-HER2 targeted therapy is received. Triple-negative breast cancer cases have significantly different (**C**) recurrence-free survival and (**D**) overall survival dependent on whether chemotherapy is received.

### Mutational Analyses

We have previously developed a bioinformatics pipeline for somatic mutation calling from breast cancer RNA-seq data [33], which has since been further implemented into the SCAN-B laboratory’s analytical routine. Within the 10-calendar year SCAN-B subset of 9,323 patients with tumor RNA-seq data, we investigated the most frequently occurring gene mutations (Figure 8). Overall and across the expressed transcriptome within this unselected and largely population-based series of early-stage breast tumor surgical specimens, each tumor was found to harbor at least 5 somatic mutations (median 44, range 5-2344). The most frequently mutated gene was *PIK3CA* (38.2%), followed by *TP53* (23.0%), and there is a long tail of less frequently mutated genes such as *MAP3K1* (9.0%), *GATA3* (7.5%), *CDH1* (7.4%), and *PTEN* (5.8%): overall 83.4% of tumors had at least 1 mutation among the top 30 most frequently mutated genes. Other mutated genes of note include *ERBB2* (4.1%), *ATM* (3.3%), *APC* (3.3%), and *ESR1* (3.1%), the last of which we have previously shown to confer de novo primary resistance to endocrine therapy [24]. When filtered to the IntOgen list of 633 driver genes, 98.2% of cases had at least one driver gene mutation (median 5, range 1-182), and 588 (92.9%) of the driver genes were mutated in at least 1 breast tumor, and 538 (85.0%) were mutated in at least 1% of tumors.

**Figure 8.**
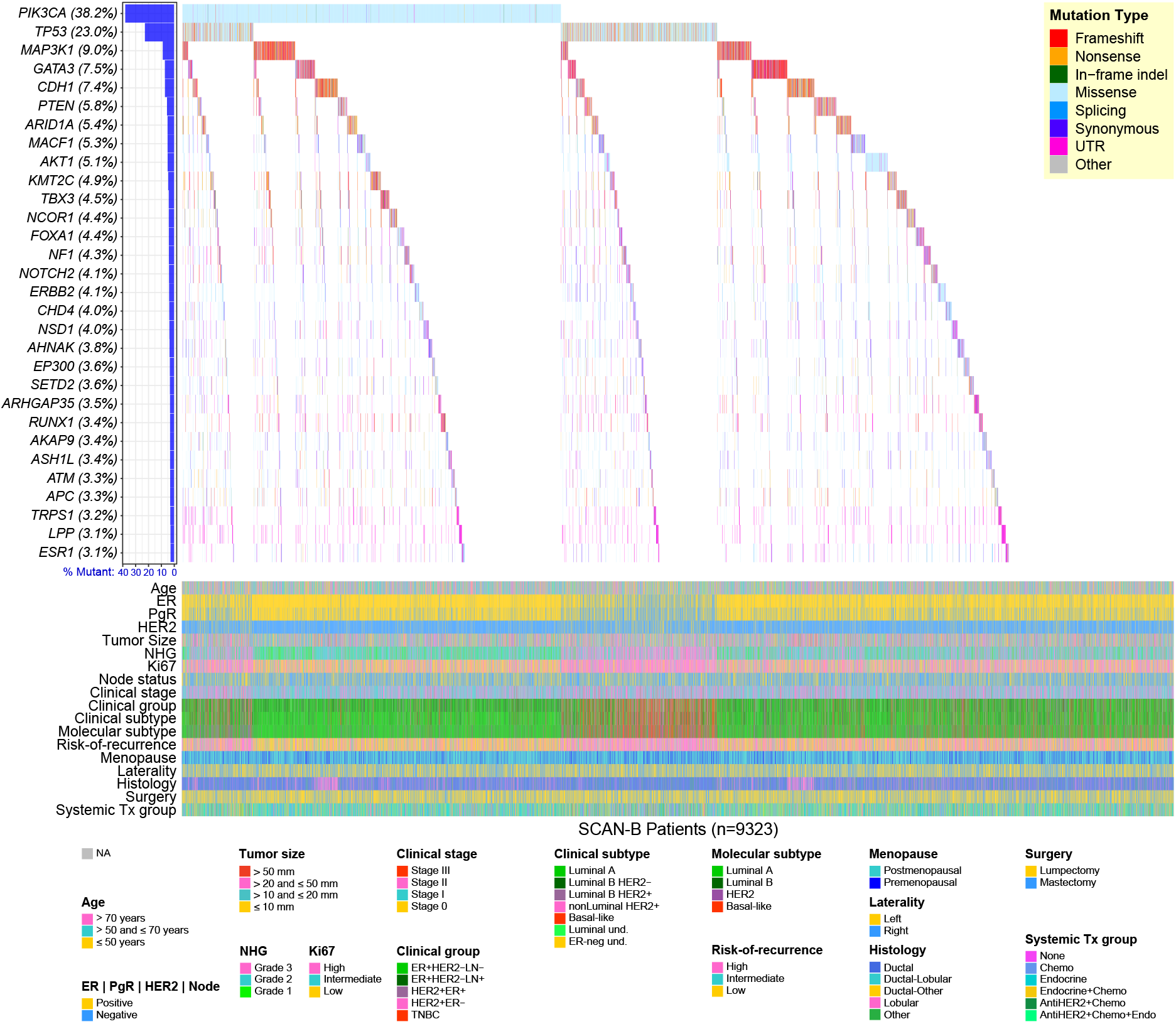
Top 30 genes with highest frequency of somatic mutations identified by RNA-sequencing. Waterfall plot of the 30 most frequently putative somatically mutated genes (rows) across the 9,323 SCAN-B tumor samples within the reported period and analyzed by RNA-seq (columns). Genes are ranked from top to bottom by mutation frequency. Samples are sorted by alteration occurrence. Mutations are colored by predicted functional impact.

### Implementation of a SCAN-B Test

In the autumn of 2021, SCAN-B reached a major milestone when it’s first developed and validated test was successfully transitioned from the research setting into the clinical setting for patients within Skåne and Blekinge, the two southernmost counties of Sweden with a population of approximately 1.42 million inhabitants and 160,000 inhabitants, respectively. Since Q4 2021, the single-sample predictor models for breast cancer molecular subtyping (SSP-PAM50 and SSP-Subtype), and risk-of-recurrence (SSP-ROR) score [41], were technology-transferred from the Lund SCAN-B laboratory into Region Skåne’s Section for Molecular Diagnostics (SMD), Laboratory Medicine, situated within the Skåne University Hospital Comprehensive Cancer Centre. This implementation was made possible in part by the experiences of the SCAN-B lab in performing RNA-seq in “real-time”, with RNA-seq data generated within approximately 1 week of surgery. A concerted effort was made by the SCAN-B lab to perform real-time analysis during the period 2015-2019, and a pilot feasibility study was performed to deliver a SCAN-B “research report” to the multidisciplinary team conference for all patients consented at Helsingborg Hospital. The Section for Clinical Pathology at the Department of Clinical Genetics, Pathology and Molecular Diagnostics, Laboratory Medicine, assisted in establishing updated routines for tissue sampling, including the 2 mm punch biopsy sample that allows for assessment of representativity of the sampled material.

Since its clinical implementation through January 2026, over 3000 post-menopausal patients with ER+ HER2– breast cancer and 0-3 positive LN have received the SSP-Subtype and SSP-ROR clinical report (Figure 1E); almost all Skåne and Blekinge patients enroll in SCAN-B as they did prior to clinical implementation. Importantly, remainder RNA/DNA/flow-through for Region Skåne and Region Blekinge patients with SCAN-B consent, after processing at SMD, are delivered to the main SCAN-B biobank. Moreover, all RNA-seq data are delivered from SMD to the SCAN-B LIMS infrastructure for the consented SCAN-B patients. Thus, the population-based nature and excellent recruitment statistics have been largely unaffected by the clinical implementation process.

## DISCUSSION

Launched in 2010, the SCAN-B initiative embarked on a mission to integrate gene expression and genomic tumor profiling into the clinical routine of breast cancer management. Over the last decade, SCAN-B aimed to enhance tumor classification and prognostication, to predict response to treatment, and reveal new tumor biology, ultimately to make these advancements accessible to future patients. To some extent, we have made significant progress to all of these aims, exemplified by more than 50 publications from the core SCAN-B team, more than 2000 citations to these papers, more than 9000 sample datasets made available in public repositories, and more than 800 search results in Google Scholar which mention SCAN-B or make use of its data (see e.g. references [23–25, 31, 33, 34, 64–70]).

Gene expression tests like Oncotype DX^®^ and Prosigna^®^ have been instrumental in guiding adjuvant chemotherapy decisions, stratifying patients by recurrence risk, and enhancing patient outcomes. In tandem, SCAN-B’s comprehensive molecular profiling using RNA-sequencing exemplifies the next step in this evolutionary trajectory. The project’s integration of SSP models into clinical decision-making represents a proof-of-concept of the circular nature of SCAN-B, where enrolled patients and their samples contribute to discovery of novel biomarker test(s), further prospectively collected patients contribute to the validation of such tests, and the same network structure can then be utilized to deliver such tests to the clinic to benefit future patients.

The inclusive enrollment strategy of SCAN-B, which encompasses all breast cancer patients at participating sites, marks a significant advancement over previous studies. Unlike other large breast cancer cohorts, which often suffer from a lack of representation due to outdated and/or heterogeneous treatments and non-population-based recruitment, SCAN-B has prospectively enrolled patients treated with modern, nationally standardized care regimens. Thus, the conclusions drawn from SCAN-B are more likely to be both representative and generalizable to a wider population. This factor is critical, as previous analyses of public breast cancer gene expression datasets have revealed a number of biases in the cohorts, thus potentially leading to less generalizable signatures and models [71, 72]. In contrast, SCAN-B’s population-based nature and basis in real-world conditions should provide a more accurate reflection of the disease spectrum at the population level.

SCAN-B’s design, which includes the collection of both tumor samples and serial blood samples, permits a dynamic observation of tumor biology and the body’s response to treatment. By analyzing these samples at predefined time points, the initiative may be able to capture a longitudinal perspective of the disease’s progression and response to therapy [73].

The commitment to real-time analysis and the biobanking of specimens linked to high-quality clinicopathological and follow-up information establish SCAN-B as a state-of-the-art model for molecular cancer research.

Looking ahead, the vast data collection and analyses facilitated by SCAN-B should continue to drive innovations in breast cancer diagnostics, prognostics, and treatment prediction. Our comprehensive approach emphasizes the critical need for molecular insights in clinical decision-making. We intend to add multi-modal analyses, some of which in turn could eventually also be implemented clinically, and have already begun within research projects with whole-genome and whole-exome sequencing, SNP chip genotyping, liquid biopsy circulating tumor cell and circulating tumor DNA monitoring (CTC/ctDNA), spatial -omics, digital pathology, and other analyses, as well as using FFPE-derived nucleic acids, and we welcome collaborations. As we continue to generate and collect data and reveal insights, SCAN-B is poised to contribute significantly to a future where breast cancer treatment is even more closely aligned with the molecular characteristics of each patient’s tumor, embodying the true essence of personalized medicine.

## Data Availability

The gene expression data derived from RNA-sequencing and microarray technologies, utilized in our prior publications, have been made publicly accessible through the NCBI Gene Expression Omnibus (GSE60789, GSE81538, GSE81540, GSE96058) and Mendeley Data repository (doi.dx.org/10.17632/yzxtxn4nmd.3). A complete deposit of all curated data summarized herein for the accrued SCAN-B early breast cancer cohort (2010-2020), together with RNA-sequencing gene expression data for the RNA-sequencing processed subset, is publicly available at Mendeley Data (doi.dx.org/10.17632/w8tsvd5zrf.1).

## ACKNOWLEDGEMENTS

The authors thank the all of patients, clinicians, and hospital staff participating in this study, the staff and former staff of the central SCAN-B laboratory and administration of the Division of Oncology, Lund University, the Swedish National Breast Cancer Quality Registry (NKBC), Regional Cancer Center South, Region Skåne Biobank, Unilabs, and the South Swedish Breast Cancer Group (SSBCG). We also thank Dorthe Grabau (deceased) for helping to initiate the project, and Joyce Carlsson for her assistance, and Sara Söderstjerna and Susanne Dieroff Hay for their engagement. SCAN-B is generously supported by the Mrs. Berta Kamprad Foundation, who were instrumental in the initiation of SCAN-B, and this work was also supported in part by the Swedish Cancer Society; Swedish Research Council; Governmental Funding of Clinical Research within National Health Service; Swedish Research Council for Health, Working Life and Welfare; Lund University Medical Faculty; Knut and Alice Wallenberg Foundation, Mats Paulsson Foundation; Lund-Lausanne L2-Bridge/Biltema Foundation; Cancera Foundation; Crafoord Foundation; Gunnar Nilsson Cancer Foundation; Krapperup Foundation; BioCARE Research Program; King Gustaf Vth Jubilee Fund; Skåne University Hospital Foundation; VINNOVA; European Commission Horizon 2020 MSCA-COFUND-2016 754299 CanFaster doctoral programme and MSCA-COFUND-2018 847583 CanFaster postdoctoral programme.

## AUTHORS’ CONTRIBUTIONS

LHS, JH, JoM, MM, JaM, JoM, CL, LR, NL, CH, ÅB, and JV-C conceived of the study. LHS, SKG-S, JH, AEd, MM, JoM, JaM, CL, LR, NL, CH, ÅB, and JV-C participated in establishing the clinical infrastructure. SKG-S, AEd, FK, EN, JoM, JaM, MR, KÅ, CI, FG, LÅ, BA, ME, MS, AC, TSv, HT, JB, LM, A-KF, A-CK, ZE, PRB, HL, TSj, MM, AEh, LR, and NL provided clinical information. LHS, JH, NN, ML, JV, and JV-C performed experiments. LHS, HD, PM, CB, SG, JH, NN, AEh, CH, and JV-C analyzed data. LHS, IH, AEd, FK, EM, P-OB, CF, CL, AEh, LR, NL, CH, ÅB, and JV-C are members of the SCAN-B Steering Committee. JoM, JaM, and MM are former members of the SCAN-B Steering Committee. LHS and HD drafted the manuscript with assistance from PM, CH, ÅB, and JV- C. All authors critically revised and approved the final manuscript.

## REFERENCES

1. Lei S, Zheng R, Zhang S, Wang S, Chen R, Sun K, et al. Global patterns of breast cancer incidence and mortality: A population-based cancer registry data analysis from 2000 to 2020. Cancer Commun (Lond). 2021;41:1183–94.

2. van’t Veer LJ, Dai H, van de Vijver MJ, He YD, Hart AAM, Mao M, et al. Gene expression profiling predicts clinical outcome of breast cancer. Nature. 2002;415:530–6.

3. Paik S, Shak S, Tang G, Kim C, Baker J, Cronin M, et al. A Multigene Assay to Predict Recurrence of Tamoxifen-Treated, Node-Negative Breast Cancer. N Engl J Med. 2004;351:2817–26.

4. Reis-Filho JS, Westbury C, Pierga JY. The impact of expression profiling on prognostic and predictive testing in breast cancer. J Clin Pathol. 2006;59:225–31.

5. Pescia C, Guerini-Rocco E, Viale G, Fusco N. Advances in Early Breast Cancer Risk Profiling: From Histopathology to Molecular Technologies. Cancers (Basel). 2023;15:5430.

6. Wallden B, Storhoff J, Nielsen T, Dowidar N, Schaper C, Ferree S, et al. Development and verification of the PAM50-based Prosigna breast cancer gene signature assay. BMC Med Genomics. 2015;8:54.

7. Parker JS, Mullins M, Cheang MCU, Leung S, Voduc D, Vickery T, et al. Supervised Risk Predictor of Breast Cancer Based on Intrinsic Subtypes. J Clin Oncol. 2009;27:1160–7.

8. Mallmann MR, Staratschek-Jox A, Rudlowski C, Braun M, Gaarz A, Wolfgarten M, et al. Prediction and prognosis: impact of gene expression profiling in personalized treatment of breast cancer patients. EPMA J. 2010;1:421–37.

9. Cardoso F, van’t Veer LJ, Bogaerts J, Slaets L, Viale G, Delaloge S, et al. 70-Gene Signature as an Aid to Treatment Decisions in Early-Stage Breast Cancer. N Engl J Med. 2016;375:717–29.

10. Loibl S, André F, Bachelot T, Barrios CH, Bergh J, Burstein HJ, et al. Early breast cancer: ESMO Clinical Practice Guideline for diagnosis, treatment and follow-up. Ann Oncol. 2024;35:159–82.

11. Ross E, Swallow J, Kerr A, Cunningham-Burley S. Online accounts of gene expression profiling in early-stage breast cancer: Interpreting genomic testing for chemotherapy decision making. Health Expect. 2019;22:74–82.

12. Sparano JA, Gray RJ, Makower DF, Pritchard KI, Albain KS, Hayes DF, et al. Adjuvant Chemotherapy Guided by a 21-Gene Expression Assay in Breast Cancer. N Engl J Med. 2018;379:111–21.

13. Stemmer SM, Steiner M, Rizel S, Soussan-Gutman L, Ben-Baruch N, Bareket-Samish A, et al. Clinical outcomes in patients with node-negative breast cancer treated based on the recurrence score results: evidence from a large prospectively designed registry. NPJ breast cancer. 2017;3:33.

14. Henry NL, Bedard PL, DeMichele A. Standard and Genomic Tools for Decision Support in Breast Cancer Treatment. Am Soc Clin Oncol Educ Book. 2017:106–15.

15. Malone ER, Oliva M, Sabatini PJB, Stockley TL, Siu LL. Molecular profiling for precision cancer therapies. Genome Med. 2020;12:8.

16. Prat A, Pineda E, Adamo B, Galván P, Fernández A, Gaba L, et al. Clinical implications of the intrinsic molecular subtypes of breast cancer. Breast. 2015;24 **Suppl 2**:S26–35.

17. Selli C, Dixon JM, Sims AH. Accurate prediction of response to endocrine therapy in breast cancer patients: current and future biomarkers. Breast Cancer Res. 2016;18:118.

18. Wang X, Collet L, Rediti M, Debien V, De Caluwé A, Venet D, et al. Predictive Biomarkers for Response to Immunotherapy in Triple Negative Breast Cancer: Promises and Challenges. J Clin Med. 2023;12:953.

19. Zardavas D, Irrthum A, Swanton C, Piccart M. Clinical management of breast cancer heterogeneity. Nat Rev Clin Oncol. 2015;12:381–94.

20. Garrido-Castro AC, Lin NU, Polyak K. Insights into Molecular Classifications of Triple-Negative Breast Cancer: Improving Patient Selection for Treatment. Cancer Discov. 2019;9:176–98.

21. van Vliet MH, Horlings HM, van de Vijver MJ, Reinders MJT, Wessels LFA. Integration of clinical and gene expression data has a synergetic effect on predicting breast cancer outcome. PloS One. 2012;7:e40358.

22. Saal LH, Vallon-Christersson J, Häkkinen J, Hegardt C, Grabau D, Winter C, et al. The Sweden Cancerome Analysis Network - Breast (SCAN-B) Initiative: a large-scale multicenter infrastructure towards implementation of breast cancer genomic analyses in the clinical routine. Genome Med. 2015;7:20.

23. Aine M, Boyaci C, Hartman J, Häkkinen J, Mitra S, Campos AB, et al. Molecular analyses of triple-negative breast cancer in the young and elderly. Breast Cancer Res. 2021;23:20.

24. Dahlgren M, George AM, Brueffer C, Gladchuk S, Chen Y, Vallon-Christersson J, et al. Preexisting Somatic Mutations of Estrogen Receptor Alpha (ESR1) in Early-Stage Primary Breast Cancer. JNCI Cancer Spectr. 2021;5:pkab028.

25. Dihge L, Vallon-Christersson J, Hegardt C, Saal LH, Häkkinen J, Larsson C, et al. Prediction of Lymph Node Metastasis in Breast Cancer by Gene Expression and Clinicopathological Models: Development and Validation within a Population-Based Cohort. Clin Cancer Res. 2019;25:6368–81.

26. Dong H, Wang S. Exploring the cancer genome in the era of next-generation sequencing. Front Med. 2012;6:48–55.

27. Larsson C, Ehinger A, Winslow S, Leandersson K, Klintman M, Dahl L, et al. Prognostic implications of the expression levels of different immunoglobulin heavy chain-encoding RNAs in early breast cancer. NPJ breast cancer. 2020;6:1–13.

28. Lundgren C, Bendahl P-O, Borg Å, Ehinger A, Hegardt C, Larsson C, et al. Agreement between molecular subtyping and surrogate subtype classification: a contemporary population-based study of ER-positive/HER2-negative primary breast cancer. Breast Cancer Res Treat. 2019;178:459–67.

29. Meyerson M, Gabriel S, Getz G. Advances in understanding cancer genomes through second-generation sequencing. Nat Rev Genet. 2010;11:685–96.

30. Rydén L, Loman N, Larsson C, Hegardt C, Vallon-Christersson J, Malmberg M, et al. Minimizing inequality in access to precision medicine in breast cancer by real-time population-based molecular analysis in the SCAN-B initiative. Br J Surg. 2018;105:e158–e68.

31. Staaf J, Glodzik D, Bosch A, Vallon-Christersson J, Reuterswärd C, Häkkinen J, et al. Whole-genome sequencing of triple-negative breast cancers in a population-based clinical study. Nat Med. 2019;25:1526–33.

32. Vallon-Christersson J, Häkkinen J, Hegardt C, Saal LH, Larsson C, Ehinger A, et al. Cross comparison and prognostic assessment of breast cancer multigene signatures in a large population-based contemporary clinical series. Sci Rep. 2019;9:12184.

33. Brueffer C, Gladchuk S, Winter C, Vallon-Christersson J, Hegardt C, Häkkinen J, et al. The mutational landscape of the SCAN-B real-world primary breast cancer transcriptome. EMBO Mol Med. 2020;12:e12118.

34. Brueffer C, Vallon-Christersson J, Grabau† D, Ehinger A, Häkkinen J, Hegardt C, et al. Clinical Value of RNA Sequencing–Based Classifiers for Prediction of the Five Conventional Breast Cancer Biomarkers: A Report From the Population-Based Multicenter Sweden Cancerome Analysis Network—Breast Initiative. JCO Precis Oncol. 2018:1–18.

35. Dalal H, Dahlgren M, Gladchuk S, Brueffer C, Gruvberger-Saal SK, Saal LH. Clinical associations of ESR2 (estrogen receptor beta) expression across thousands of primary breast tumors. Sci Rep. 2022;12:4696.

36. Gruvberger S, Ringnér M, Chen Y, Panavally S, Saal LH, Borg A n, et al. Estrogen receptor status in breast cancer is associated with remarkably distinct gene expression patterns. Cancer Res. 2001;61:5979–84.

37. Hedenfalk I, Duggan D, Chen Y, Radmacher M, Bittner M, Simon R, et al. Gene-expression profiles in hereditary breast cancer. N Engl J Med. 2001;344:539–48.

38. Jönsson G, Staaf J, Vallon-Christersson J, Ringnér M, Gruvberger-Saal SK, Saal LH, et al. The retinoblastoma gene undergoes rearrangements in BRCA1-deficient basal-like breast cancer. Cancer Res. 2012;72:4028–36.

39. Meng P, Dalal H, Chen Y, Brueffer C, Gladchuk S, Alcaide M, et al. Digital PCR quantification of ultrahigh ERBB2 copy number identifies poor breast cancer survival after trastuzumab. NPJ breast cancer. 2024;10:1–10.

40. Saal LH, Johansson P, Holm K, Gruvberger-Saal SK, She Q-B, Maurer M, et al. Poor prognosis in carcinoma is associated with a gene expression signature of aberrant PTEN tumor suppressor pathway activity. Proc Natl Acad Sci U S A. 2007;104:7564–9.

41. Staaf J, Häkkinen J, Hegardt C, Saal LH, Kimbung S, Hedenfalk I, et al. RNA sequencing-based single sample predictors of molecular subtype and risk of recurrence for clinical assessment of early-stage breast cancer. NPJ breast cancer. 2022;8:1–17.

42. Staaf J, Ringnér M, Vallon-Christersson J, Jönsson G, Bendahl P-O, Holm K, et al. Identification of subtypes in human epidermal growth factor receptor 2--positive breast cancer reveals a gene signature prognostic of outcome. J Clin Oncol. 2010;28:1813–20.

43. Veerla S, Hohmann L, Nacer DF, Vallon-Christersson J, Staaf J. Perturbation and stability of PAM50 subtyping in population-based primary invasive breast cancer. NPJ breast cancer. 2023;9:83.

44. Lofgren L, Eloranta S, Krawiec K, Asterkvist A, Lonnqvist C, Sandelin K, et al. Validation of data quality in the Swedish National Register for Breast Cancer. BMC Public Health. 2019;19:495.

45. Häkkinen J, Nordborg N, Månsson O, Vallon-Christersson J. Implementation of an Open Source Software solution for Laboratory Information Management and automated RNAseq data analysis in a large-scale Cancer Genomics initiative using BASE with extension package Reggie. bioRxiv. 2016.

46. Saal LH, Troein C, Vallon-Christersson J, Gruvberger S, Borg Å, Peterson C. BioArray Software Environment (BASE): a platform for comprehensive management and analysis of microarray data. Genome Biol. 2002;3:software0003.1.

47. Troein C, Vallon-Christersson J, Saal LH. An introduction to BioArray Software Environment. Methods Enzymol. 2006;411:99–119.

48. Vallon-Christersson J, Nordborg N, Svensson M, Häkkinen J. BASE--2nd generation software for microarray data management and analysis. BMC bioinformatics. 2009;10:330.

49. Nalpas NC, Park SDE, Magee DA, Taraktsoglou M, Browne JA, Conlon KM, et al. Whole-transcriptome, high-throughput RNA sequence analysis of the bovine macrophage response to Mycobacterium bovis infection in vitro. BMC Genomics. 2013;14:230.

50. Parkhomchuk D, Borodina T, Amstislavskiy V, Banaru M, Hallen L, Krobitsch S, et al. Transcriptome analysis by strand-specific sequencing of complementary DNA. Nucleic Acids Res. 2009;37:e123.

51. Rydén S, Fernö M, Borg A, Hafström L, Möller T, Norgren A. Prognostic significance of estrogen and progesterone receptors in stage II breast cancer. J Surg Oncol. 1988;37:221–6.

52. Rydén S, Fernö M, Möller T, Aspegren K, Bergljung L, Killander D, et al. Long-term effects of adjuvant tamoxifen and/or radiotherapy. The South Sweden Breast Cancer Trial. Acta Oncol. 1992;31:271–4.

53. Ryden S, Möller T, Hafström L, Ranstam J, Westrup C, Wiklander O. Adjuvant therapy of breast cancer: compliance and data validity in a multicenter trial. Controlled Clin Trials. 1986;7:290–305.

54. Tennvall-Nittby L, Tengrup I, Landberg T. The total incidence of loco-regional recurrence in a randomized trial of breast cancer TNM stage II. The South Sweden Breast Cancer Trial. Acta Oncol. 1993;32:641–6.

55. Fernö M, Borg A, Johansson U, Norgren A, Olsson H, Rydén S, et al. Estrogen and progesterone receptor analyses in more than 4,000 human breast cancer samples. A study with special reference to age at diagnosis and stability of analyses. Southern Swedish Breast Cancer Study Group. Acta Oncol. 1990;29:129–35.

56. Sigurdsson H, Baldetorp B, Borg A, Dalberg M, Fernö M, Killander D, et al. Indicators of prognosis in node-negative breast cancer. N Engl J Med. 1990;322:1045–53.

57. Borg A, Haile RW, Malone KE, Capanu M, Diep A, Törngren T, et al. Characterization of BRCA1 and BRCA2 deleterious mutations and variants of unknown clinical significance in unilateral and bilateral breast cancer: the WECARE study. Hum Mutat. 2010;31:E1200–40.

58. Johannsson O, Ostermeyer EA, Håkansson S, Friedman LS, Johansson U, Sellberg G, et al. Founding BRCA1 mutations in hereditary breast and ovarian cancer in southern Sweden. Am J Clin Genet. 1996;58:441–50.

59. Loman N, Bladström A, Johannsson O, Borg A, Olsson H. Cancer incidence in relatives of a population-based set of cases of early-onset breast cancer with a known BRCA1 and BRCA2 mutation status. Breast Cancer Res. 2003;5:R175–86.

60. Loman N, Johannsson O, Bendahl PO, Borg A, Fernö M, Olsson H. Steroid receptors in hereditary breast carcinomas associated with BRCA1 or BRCA2 mutations or unknown susceptibility genes. Cancer. 1998;83:310–9.

61. Nilsson MP, Törngren T, Henriksson K, Kristoffersson U, Kvist A, Silfverberg B, et al. BRCAsearch: written pre-test information and BRCA1/2 germline mutation testing in unselected patients with newly diagnosed breast cancer. Breast Cancer Res Treat. 2018;168:117–26.

62. Jönsson G, Naylor TL, Vallon-Christersson J, Staaf J, Huang J, Ward MR, et al. Distinct genomic profiles in hereditary breast tumors identified by array-based comparative genomic hybridization. Cancer Res. 2005;65:7612–21.

63. Khan J, Saal LH, Bittner ML, Chen Y, Trent JM, Meltzer PS. Expression profiling in cancer using cDNA microarrays. Electrophoresis. 1999;20:223–9.

64. Bjorklund SS, Aure MR, Hakkinen J, Vallon-Christersson J, Kumar S, Evensen KB, et al. Subtype and cell type specific expression of lncRNAs provide insight into breast cancer. Commun Biol. 2022;5:834.

65. Demircan K, Bengtsson Y, Sun Q, Brange A, Vallon-Christersson J, Rijntjes E, et al. Serum selenium, selenoprotein P and glutathione peroxidase 3 as predictors of mortality and recurrence following breast cancer diagnosis: A multicentre cohort study. Redox Biol. 2021;47:102145.

66. Martin M, Stecklein SR, Gluz O, Villacampa G, Monte-Millan M, Nitz U, et al. TNBC-DX genomic test in early-stage triple-negative breast cancer treated with neoadjuvant taxane-based therapy. Ann Oncol. 2025;36:158–71.

67. Rediti M, Venet D, Joaquin Garcia A, Maetens M, Vincent D, Majjaj S, et al. Identification of HER2-positive breast cancer molecular subtypes with potential clinical implications in the ALTTO clinical trial. Nat Commun. 2024;15:10402.

68. Søkilde R, Persson H, Ehinger A, Pirona AC, Fernö M, Hegardt C, et al. Refinement of breast cancer molecular classification by miRNA expression profiles. BMC Genomics. 2019;20:503.

69. Villacampa G, Pascual T, Braso-Maristany F, Pare L, Martinez-Saez O, Cortes J, et al. Prognostic value of HER2DX in early-stage HER2-positive breast cancer: a comprehensive analysis of 757 patients in the Sweden Cancerome Analysis Network-Breast dataset (SCAN-B). ESMO Open. 2024;9:102388.

70. Winter C, Nilsson MP, Olsson E, George AM, Chen Y, Kvist A, et al. Targeted sequencing of BRCA1 and BRCA2 across a large unselected breast cancer cohort suggests that one-third of mutations are somatic. Ann Oncol. 2016;27:1532–8.

71. Sims AH, Smethurst GJ, Hey Y, Okoniewski MJ, Pepper SD, Howell A, et al. The removal of multiplicative, systematic bias allows integration of breast cancer gene expression datasets – improving meta-analysis and prediction of prognosis. BMC Med Genomics. 2008;1:42.

72. Xie Y, Davis Lynn BC, Moir N, Cameron DA, Figueroa JD, Sims AH. Breast cancer gene expression datasets do not reflect the disease at the population level. NPJ breast cancer. 2020;6:39.

73. Olsson E, Winter C, George A, Chen Y, Howlin J, Tang MHE, et al. Serial monitoring of circulating tumor DNA in patients with primary breast cancer for detection of occult metastatic disease. EMBO Mol Med. 2015;7:1034–47.

