## Supplementary Figure 1 for "Population-scale integration of tumor transcriptomics into breast cancer care: a decade of the SCAN-B initiative"

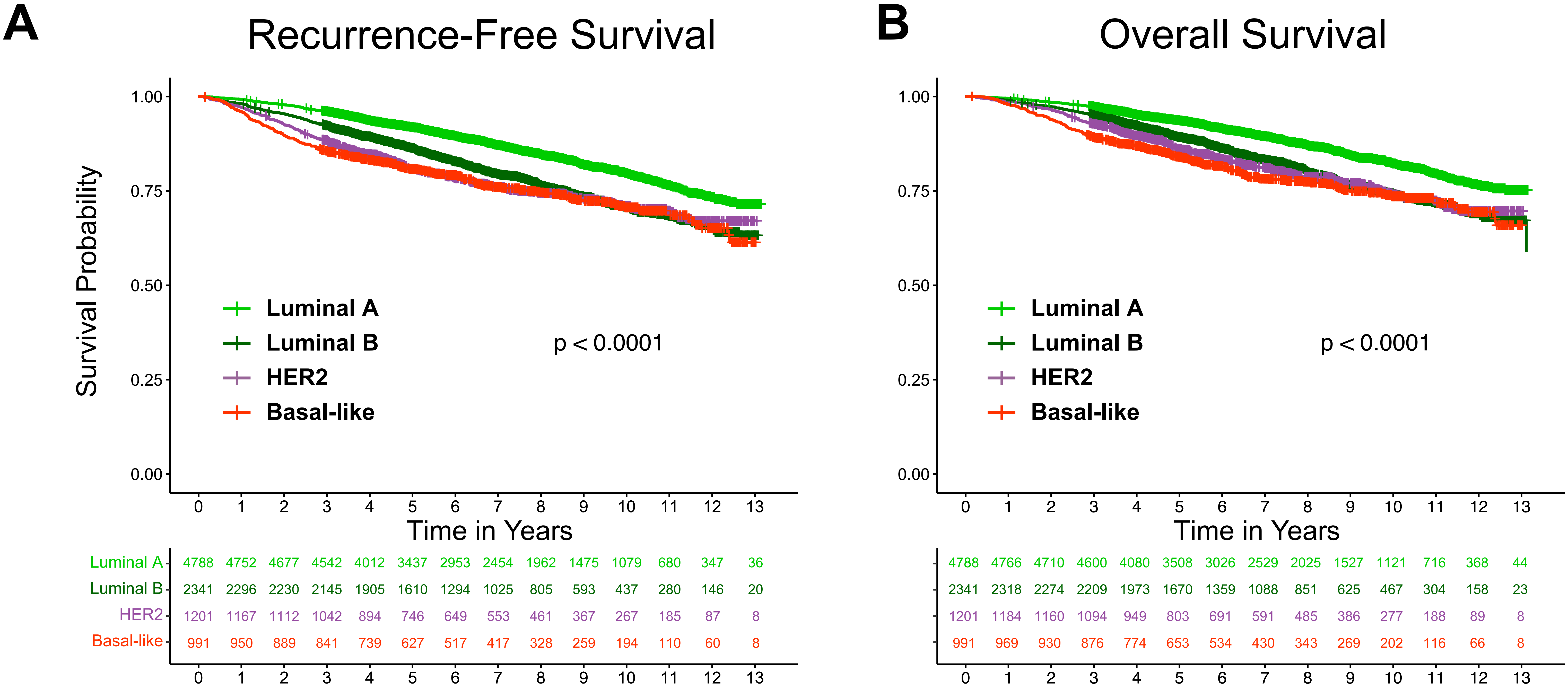


**Supplementary Figure 1**. Analyses of SCAN-B patient outcomes by molecular subtype. Kaplan-Meier survival estimates for SCAN-B patients within the reported period and with available RNA-sequencing-based molecular subtype and follow-up information for recurrence-free survival (A) and overall survival (B).
